# Long-term duration of protection of ancestral-strain monovalent vaccines and effectiveness of the bivalent BA.1 boosters against COVID-19 hospitalisation during a period of BA.5, BQ.1, CH.1.1. and XBB.1.5 circulation in England

**DOI:** 10.1101/2023.03.31.23288018

**Authors:** Freja Cordelia Møller Kirsebom, Nick Andrews, Julia Stowe, Mary Ramsay, Jamie Lopez Bernal

## Abstract

**Background:** Bivalent BA.1 booster vaccines were offered to adults aged 50 years and older and clinically vulnerable individuals as part of the autumn COVID-19 booster vaccination programme 2022 in England.

**Methods:** A test-negative case-control study was used to estimate the duration of protection of the monovalent vaccines against hospitalisation as compared to those unvaccinated. In addition, the incremental VE of the bivalent BA.1 booster vaccines was estimated relative to those with waned immunity where the last dose was at least 6 months prior amongst those aged 50 years and older.

**Findings:** The protection conferred by the monovalent vaccines was well maintained long-term: absolute VE against hospitalisation amongst those aged 65 years and older who had received at least 3 doses plateaued from 6 months after the last dose at around 50%. Incremental VE (in addition to the protection from earlier vaccines) of the bivalent BA.1 boosters against hospitalisation peaked at 53.0% (95% C.I.; 47.9-57.5%) (equivalent to an absolute VE of approximately 75%) before waning to around 35.9% (95% C.I.; 31.4-40.1%) after 10 or more weeks.

**Interpretation:** This study provides evidence of the long-term duration of protection of the monovalent vaccines, suggesting individuals at lower risk of severe disease who did not receive a booster in autumn 2022 may not require regular re-vaccination. Furthermore, this study finds good evidence that the bivalent BA.1 booster vaccines are highly effective against hospitalisation amongst those aged 50 years and older with the sub-lineages of Omicron present in the autumn/winter of 2022 in England.

**Funding:** None.

## Introduction

Monovalent vaccines developed to target the ancestral strain of SARS-CoV-2 have proven highly effective at protecting against severe COVID-19 disease (1-4). Nonetheless, the emergence of Omicron with immune-evading properties has reduced vaccine effectiveness (VE) and the longevity of the protection conferred by the vaccines (3, 4).

The first Omicron sub-lineage to emerge in the UK was BA.1 in November 2021 (5), followed by BA.2 (6) and BA.4 and BA.5 (7). In the autumn/winter of 2022/23 BA.5 dominated in September. The prevalence of BQ.1/BQ.1.1 increased from October while the prevalence of CH.1.1 and XBB.1.5 increased from December 2022 and January 2023, respectively (8). These sub-lineages have all acquired different combinations of mutations in the spike protein as compared to BA.1 (9, 10). Neutralizing of BQ.1, CH.1.1 and XBB.1.5 by sera from vaccinated and previously infected individuals was markedly reduced from that observed against BA.1 (9-14).

Many countries have opted for frequent booster programmes to maintain a high level of protection in the most vulnerable populations. Concurrently, bivalent vaccines have been developed which target both the spike proteins from ancestral wild-type SARS-CoV-2 and from immune-evading Omicron sub-lineages. Laboratory studies have found antibodies from those vaccinated with bivalent vaccines targeting the ancestral and BA.1 had increased neutralization capacity against BA.4, BA.5, and BA.2.75 compared to those vaccinated with the ancestral only vaccine (15-17).

As part of the UK COVID-19 vaccination programme, all adults in England were offered a primary course consisting of two doses of either Oxford/AstraZeneca (ChAdOx1-S), Pfizer BioNTech (Original Comirnaty®) or Moderna (Spikevax®) and an mRNA booster vaccine of either Pfizer BioNTech (Original Comirnaty®) or Moderna (Spikevax®). The last booster offered to all adults was available from 29 November 2021 (18). Subsequently, the Joint Committee on Vaccination and immunisation (JCVI) recommended an additional spring booster in 2022 for those aged 75 years and over, residents in a care home for older adults and the immunosuppressed (19). An autumn 2022 booster programme commencing 5^th^ September 2022 was recommended by the JCVI (20) and bivalent BA.1 boosters with either Pfizer BioNTech (Original/Omicron BA.1 Comirnaty®) or Moderna (Spikevax® bivalent Original/Omicron BA.1 vaccine) were offered to all adults aged 50 years and over and vulnerable individuals (21).

Here, we estimate the long-term duration of protection from the monovalent vaccines given as part of a primary course or earlier booster campaign using a test-negative case control (TNCC) study design. Additionally, we estimated the incremental vaccine effectiveness (iVE), (often also called relative VE (22)), of the bivalent BA.1 boosters against hospitalisation. The iVE of the bivalent boosters was estimated within those who had received at least two prior doses, with the last dose being at least 6 months prior, as the baseline comparator group.

## Methods

### Study design

To estimate VE against hospitalisation, a TNCC study design was used where positive PCR tests from individuals hospitalised with a respiratory code in the primary diagnosis field are cases while negative tests from such individuals are controls, as previously described (1-4).

### Data Sources

#### COVID-19 Testing Data

Pillar 1 SARS-CoV-2 PCR testing was previously available for those with a clinical need and for health and care workers. Since September 2022, Pillar 1 PCR testing has been restricted to those with respiratory disease in hospital settings. The study period for tests contributing to the assessment of previous doses was from 13^th^ June 2022 to 25^th^ December 2022, and for the assessment of bivalent booster doses from 5^th^ September 2022 to 5^th^ February 2023.

All Pillar 1 positive and negative PCR tests were extracted for the study periods of interest. Negative tests taken within 7 days of a previous negative test were dropped as these likely represent the same episode. Negative tests taken within 21 days of a subsequent positive test were also excluded as chances are high that these are false negatives. Tests within 90 days of a previous positive test were also excluded. The date of an individual’s most recent prior positive test was identified from all testing data. Individuals contributed a maximum of one negative control test (selected at random) in each period of 13^th^ June to 4^th^ September 2022, and 5^th^ September 2022 onwards. Variant status was classified based on whole genome sequencing and genotyping. Any non-Omicron cases were excluded.

#### National Immunisation Management System (NIMS)

Testing data were linked to NIMS using combinations of the unique individual NHS number, date of birth, surname, first name, and postcode using deterministic linkage. NIMS was accessed for dates of vaccination and manufacturer, sex, date of birth, ethnicity, and residential address. Addresses were used to determine index of multiple deprivation quintile. Data on risk group status (those identified as at risk previously in the pandemic and those identified recently as requiring an autumn booster by NHS CaaS (Cohorting as a Service) (23)), clinically extremely vulnerable status, severely immunosuppressed and health/social care worker status were also extracted from the NIMS. Bivalent doses given as part of the autumn booster programme were classified based on SNOMED coding and timing (administered after 5^th^ September 2022).

To estimate long-term duration of the monovalent vaccines the following individuals were excluded: those who had only a single dose, those who received a bivalent booster vaccine by their test date and those whose last dose prior to 5^th^ September 2022 was by a manufacturer other than AstraZeneca, Pfizer or Moderna.

For the analysis of the bivalent vaccines the following individuals were excluded: those who were unvaccinated, those who had received only one dose, those who received trial doses, those who received an autumn dose without receiving at least two other doses prior to 5^th^ September, those who received an autumn booster less than 12 weeks after their next most recent dose, those who received two autumn doses, those who received a vaccine coded as bivalent prior to 5^th^ September, those who received an autumn dose not coded as bivalent, and those whose last dose prior to 5^th^ September 2022 was by a manufacturer other than AstraZeneca, Pfizer or Moderna.

#### Hospital Admission Data

Hospital inpatient admissions for a range of acute respiratory illnesses (ARI) were identified from the Secondary Uses Service (SUS) (24) and were linked to the testing data using NHS number and date of birth. Admissions with an ICD-10 coded acute respiratory illness (ARI) discharge diagnosis in any diagnosis field were identified where the date of test was 1 day before and up to 2 days after the admission. Data were restricted to those with ARI in the first diagnosis field (Supplementary Table 1) and where the length of stay was at least 2 days. To estimate VE against the most severe respiratory disease outcome, tests were restricted to hospitalisations which required either supplemental oxygen (O2), treatment on an intensive care unit (ICU) or mechanical ventilation (4).

#### Covariates and adjustment

Potential confounding variables were week of test date, gender, age (five year age bands), risk group status, care home status, health and social care worker status, region, IMD quintile, ethnicity and the likely variant of an individual’s most recent prior infection.

### Statistical methods

Multivariable logistic regression was used with the test result as the outcome, vaccination status as the primary variable of interest and with confounder adjustment as described above. VE was calculated as 1-odds ratio and given as a percentage.

To estimate long term waning of doses given prior to 5^th^ September 2022 intervals used were 0 to 1 week, 2 weeks to 2 months, 3 to 5 months, 6 to 8 months, 9 to 11 months and 12 to 14 months and 15 or more months after a second dose and after a third or fourth dose. Note that very few will have received second doses recently in the study period. The analysis was split by those aged 18 to 64 years and those aged 65 years and older. The absolute VE (aVE) was estimated with the baseline comparator group being the unvaccinated. The iVE of the fourth dose was also estimated relative to a third dose given at least 6 months prior to sample date within those aged 75 years and older (the target age for this dose).

Incremental VE of the bivalent booster was estimated amongst those who had received at least two doses prior to the 5^th^ September 2022 and whose final dose prior to the 5^th^ September 2022 was at least 6 months before their test date, with those who did not receive a bivalent booster being the comparator group. VE was estimated at the following intervals since booster vaccination; 0 to 6 days, 7 to 13 days, 2 to 4 weeks, 5 to 9 weeks and 10 or more weeks. Primary analyses were restricted to those aged 50 years and older and estimated by manufacturers combined as well as stratified by manufacturer.

Stratified analyses were conducted to estimate iVE of the bivalent boosters for the following intervals between the final dose received prior to 5^th^ September and the test date; 6 weeks to 5 months, 6 to 8 months, 9 to 11 months and 12 or more months. Sensitivity analyses also investigated iVE of the bivalent booster in those who had received at least 3 and at least 4 doses prior to the 5^th^ September. Additionally, analyses were done restricting to those aged 75 years and older who received at least 4 doses prior to 5^th^ September with a 6 to 8 month interval to the last dose since 5^th^ September, and restricting to those aged 65 years and older who received at least two doses prior to 5^th^ September with a 6 to 8 month interval to the last dose since 5^th^ September.

Analyses were also conducted to remove the adjustment for previous positivity and to estimate iVE against hospitalisation regardless of ICD-10 coding in the primary diagnosis field.

## Results

### Duration of protection of the monovalent vaccines against hospitalisation in those who did not receive a bivalent booster

There were 63,251 eligible tests from hospitalised individuals aged 18 years and older, with 19,841 being cases and 43,410 being controls. Full descriptive characteristics are available in Supplementary Table 2 and Supplementary Figure 1. The duration of protection was estimated amongst those who had received two doses and amongst those who had received at least three doses, stratified by those aged 18 to 64 years and those aged 65 years and older (Table 1, Figure 1).

**Table 1.**
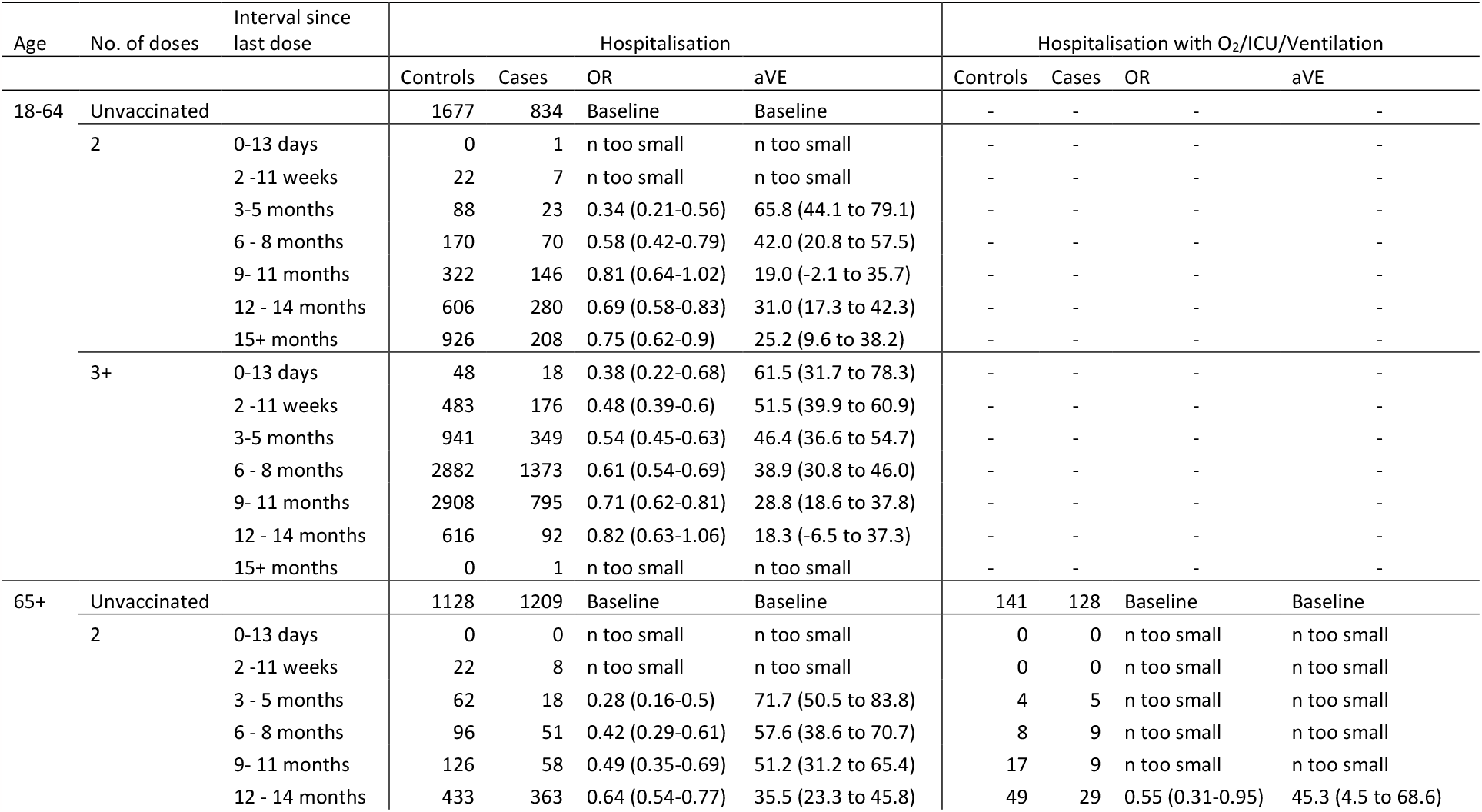
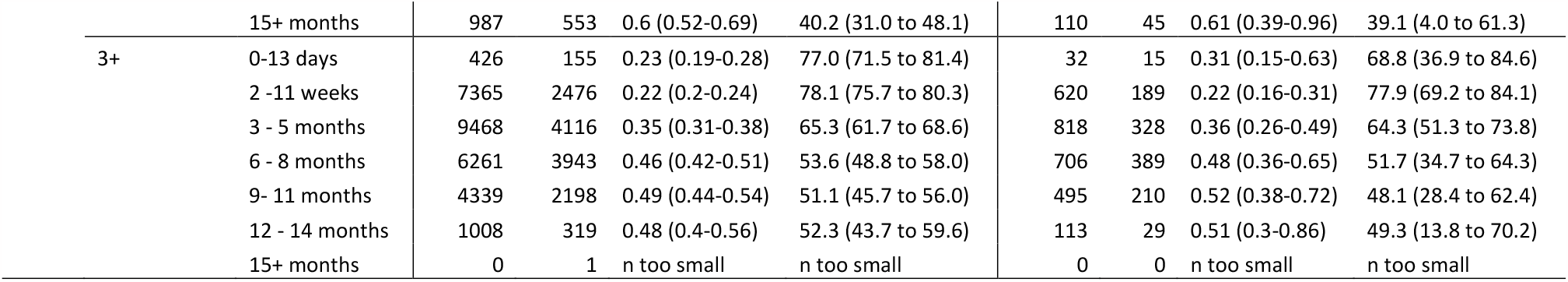
Absolute vaccine effectiveness (aVE) against hospitalisation of a second or at least a third dose of the monovalent vaccines amongst those who did not receive at bivalent BA.1 vaccine.

**Table 2.**
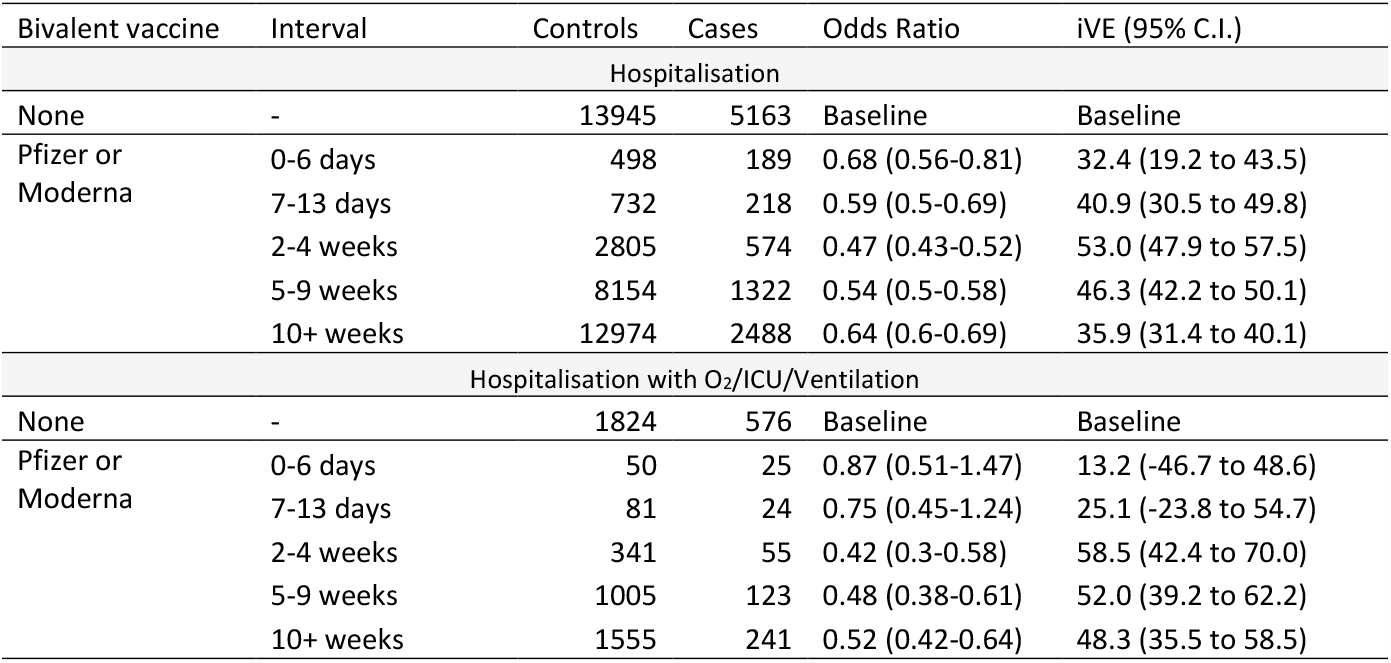
Incremental vaccine effectiveness (iVE) against hospitalisation of the bivalent booster amongst those aged 50 years and older in England. Bivalent booster manufacturers (Pfizer and Moderna) combined.

**Figure 1.**
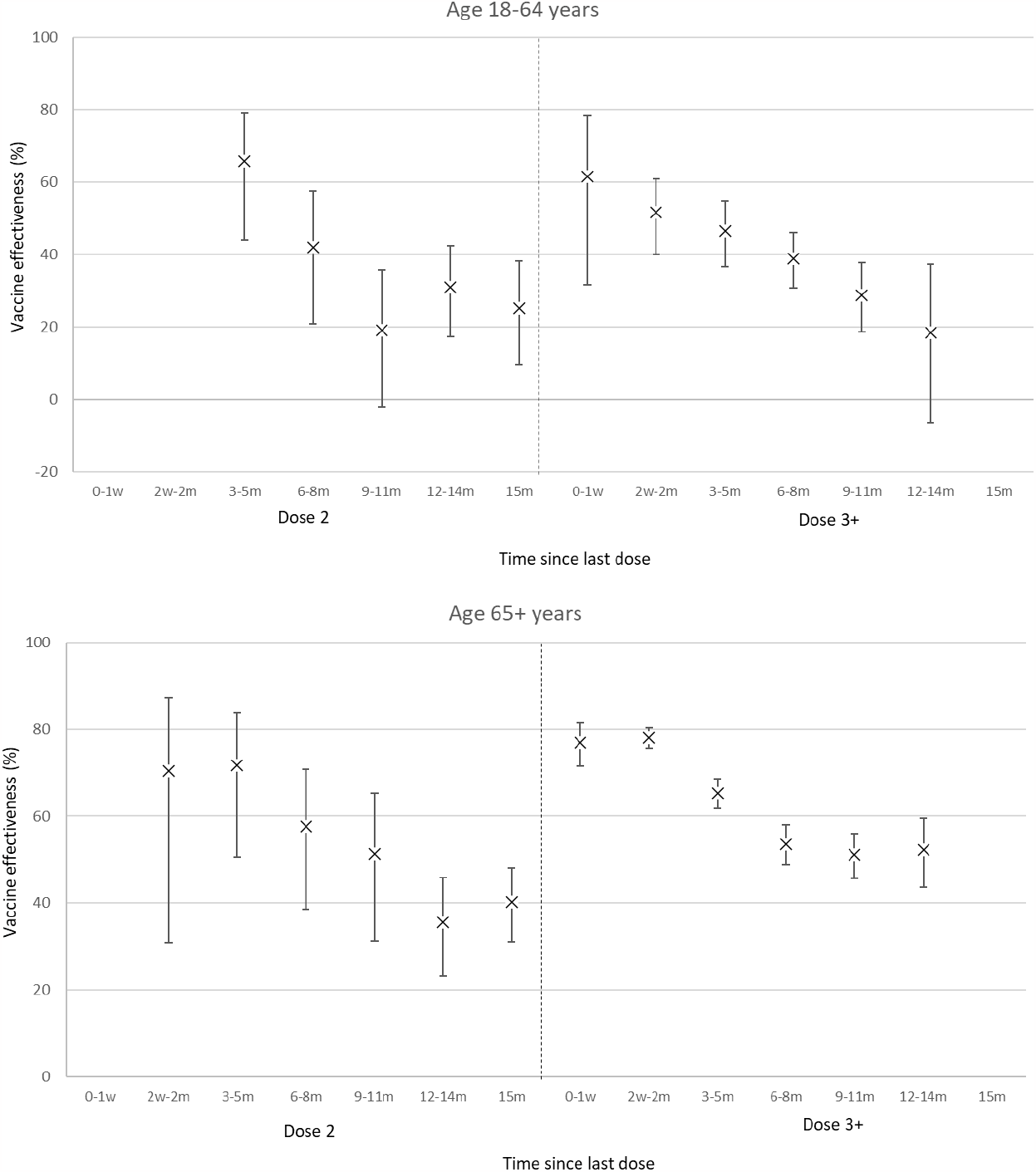
Absolute vaccine effectiveness (aVE) against hospitalisation of a second or at least a third dose of the monovalent vaccines amongst those who did not receive a bivalent BA.1 vaccine. The baseline comparator group is the unvaccinated.

Amongst those aged 18 to 64 years, the aVE of two doses was 65.8% (95% C.I.; 44.1 to 79.1%) after 3 to 5 months (Table 1, Figure 1), this waned to 25.2% (95% C.I.; 9.6 to 38.2%) after 15 or more months. For those who had received at least three doses, aVE was 46.4% (95% C.I.; 36.6 to 54.7%) after 3 to 5 months, and 18.3% (95% C.I.; -6.5 to 37.3%) after 12 to 14 months. Amongst those aged 65 years and older, aVE after a second dose was 71.7% (95% C.I.; 50.5 to 83.8%) after 3 to 5 months, plateauing after approximately 12 months; aVE was 40.2% (95% C.I.; 31.0 to 48.1%) after 15 or more months. Amongst those who had received at least three doses, the protection conferred by their most recent dose was 65.3% (95% C.I.; 61.7 to 68.6%) after 3 to 5 months. Protection after at least 3 doses plateaued after approximately 6 months, aVE was 52.3% (95% C.I.; 43.7 to 59.6%) after 12 to 14 months. Removing the adjustment for previous positivity did not affect VE estimates (Supplementary Table 3). Absolute VE against the most severe disease requiring hospitalisation with supplemental oxygen (O2), ICU or mechanical ventilation was 39.1% (95% C.I.; 4.0 to 61.3%) after 15 or more months after two doses, and 49.3% (95% C.I.; 13.8 to 70.2%) after 12 to 14 months after three or more doses.

### Duration of protection of the monovalent spring booster against hospitalisation in those aged 75 years and older

Long term waning of the most recent monovalent dose where the dose number was at least a fourth dose was also estimated amongst those aged 75 years and older (Supplementary Table 4). Here, the iVE was estimated relative to those who had received a third dose. Protection peaked at around 50%, before waning to a similar level to the baseline protection conferred by a waned third dose (hence no iVE).

### Bivalent BA.1 booster incremental vaccine effectiveness against hospitalisation

There were 49,062 eligible tests from hospitalised individuals aged 50 years and older, with 9,954 being cases and 39,108 being controls. Full descriptive characteristics are available in Supplementary Table 5 and Supplementary Figure 2.

The iVE of the bivalent BA.1 boosters peaked after 2 to 4 weeks at 53.0% (95% C.I.; 47.9 to 57.5%) and waned to 35.9% (95% C.I.; 31.4 to 40.1%) after 10 or more weeks (Table 2, Supplementary Figure 3). When stratified by manufacturer, there was no significant difference between Pfizer and Moderna boosters (Table 3, Figure 2a). The iVE of the Pfizer booster peaked at 2 to 4 weeks at 47.2% (95% C.I.; 39.4 to 54.1%), before waning to 38.0% (95% C.I.; 31.0 to 44.3%) after 10 or more weeks. The iVE of the Moderna booster peaked at 57.8% (95% C.I.; 51.6 to 63.3%) after 2 to 4 weeks and was 34.1% (95% C.I.; 29.2 to 38.7%) after 10 or more weeks. The iVE against severe hospitalisation peaked at 60.9% (95% C.I.; 44.1 to 72.6%) and 69.3% (95% C.I.; 51.4 to 80.6%) for the Pfizer and Moderna bivalent boosters, respectively (Table 3, Figure 2b).

**Table 3.** Incremental vaccine effectiveness (iVE) against hospitalisation and against hospitalisation requiring supplemental oxygen (O_2_), treatment in an intensive care unit (ICU) or mechanical ventilation amongst those aged 50 years and older in England, stratified by bivalent booster manufacturer.

| Bivalent vaccine | Interval | Controls | Cases | Odds Ratio | iVE (95% C.I.) |
| --- | --- | --- | --- | --- | --- |
| Hospitalisation |  |  |  |  |  |
| None | - | 13945 | 5163 | Baseline | Baseline |
| Pfizer | 0-6 days | 303 | 105 | 0.74 (0.59-0.94) | 25.7 (5.9 to 41.3) |
|  | 7-13 days | 463 | 118 | 0.68 (0.55-0.84) | 31.8 (15.5 to 44.9) |
|  | 2-4 weeks | 1649 | 268 | 0.53 (0.46-0.61) | 47.2 (39.4 to 54.1) |
|  | 5-9 weeks | 4394 | 678 | 0.56 (0.51-0.62) | 43.7 (38.1 to 48.8) |
|  | 10+ weeks | 3086 | 575 | 0.62 (0.56-0.69) | 38.0 (31.0 to 44.3) |
| Moderna | 0-6 days | 195 | 84 | 0.59 (0.45-0.77) | 41.3 (23.4 to 55.1) |
|  | 7-13 days | 269 | 100 | 0.49 (0.39-0.62) | 51.0 (37.6 to 61.5) |
|  | 2-4 weeks | 1156 | 306 | 0.42 (0.37-0.48) | 57.8 (51.6 to 63.3) |
|  | 5-9 weeks | 3760 | 644 | 0.52 (0.48-0.58) | 47.5 (42.1 to 52.4) |
|  | 10+ weeks | 9888 | 1913 | 0.66 (0.61-0.71) | 34.1 (29.2 to 38.7) |
| Hospitalisation with O<sub>2</sub>/ICU/Ventilation |  |  |  |  |  |
| None | - | 1824 | 576 | Baseline | Baseline |
| Pfizer | 0-6 days | 34 | 11 | 0.65 (0.32-1.34) | 35.1 (-33.6 to 68.4) |
|  | 7-13 days | 53 | 14 | 0.92 (0.49-1.75) | 7.6 (-75.2 to 51.2) |
|  | 2-4 weeks | 202 | 29 | 0.53 (0.35-0.81) | 46.6 (18.8 to 64.9) |
|  | 5-9 weeks | 549 | 68 | 0.51 (0.38-0.68) | 49.5 (32.3 to 62.3) |
|  | 10+ weeks | 375 | 46 | 0.39 (0.27-0.56) | 60.9 (44.1 to 72.6) |
| Moderna | 0-6 days | 16 | 14 | n too small | n too small |
|  | 7-13 days | 28 | 10 | n too small | n too small |
|  | 2-4 weeks | 139 | 26 | 0.31 (0.19-0.49) | 69.3 (51.4 to 80.6) |
|  | 5-9 weeks | 456 | 55 | 0.44 (0.32-0.6) | 56.4 (39.8 to 68.4) |
|  | 10+ weeks | 1180 | 195 | 0.55 (0.44-0.68) | 45.2 (31.6 to 56.0) |

**Figure 2.**
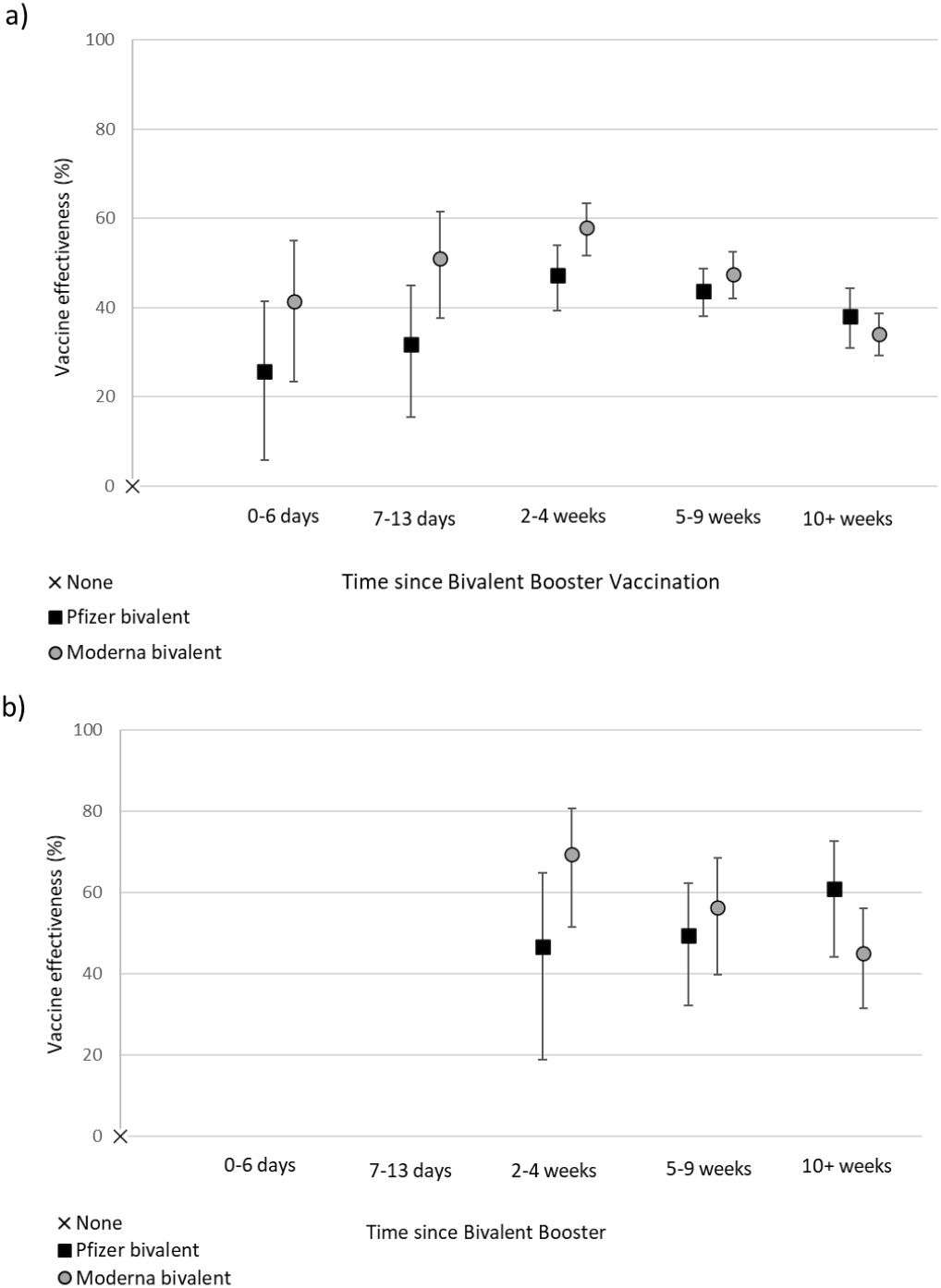
Incremental vaccine effectiveness (iVE) against (a) hospitalisation and (b) hospitalisation requiring supplemental oxygen, treatment on intensive care unit or mechanical ventilation amongst those aged 50 years and older. Bivalent booster split by manufacturer. The baseline comparator group is those who received at least two doses, and their last dose at least 6 months ago.

The iVE of the bivalent boosters increased as the time since the most recent prior dose increased; with a 6 week to 5 month interval to the most recent dose, the iVE was 46.0% (95% C.I.; 36.8 to 53.8%) after 2 to 4 weeks (Supplementary Table 6). With a 6 to 8 month interval to the most recent dose, the iVE was 46.5% (95% C.I.; 37.9 to 53.9%) whilst with an interval of 9 to 11 months the incremental VE of the bivalent boosters increased to 59.9% (95% C.I.; 50.5 to 67.5%) and to 55.4% (95% C.I.; 48.7 to 61.2%) where the interval was at least 12 months (Supplementary Table 6).

Removing the adjustment for previous positivity had minimal impact on the iVE of the bivalent BA.1 boosters, as did including all hospitalisations regardless of ICD-10 coding (Supplementary Table 6). There was no difference in the iVE of the bivalent boosters between those who had had at least two, three or four prior doses. The iVE was estimated in those aged 75 years and older who had received at least 4 doses prior to the autumn programme to investigate the added benefit of the bivalent booster doses in those who likely received a spring booster. In this group, the iVE of the bivalent BA.1 boosters was similar to that observed previously at 48.0% (95% C.I.; 38.8 to 55.7%) after 2 to 4 weeks.

## Discussion

We estimated the duration of protection of the monovalent vaccines against severe disease in adults who had not received a bivalent BA.1 booster and found that protection plateaus around 6 months after the most recent dose and is long lasting, with a modest level of protection observed even after 15 or more months. We also estimated the iVE of the bivalent BA.1 boosters against hospitalisation and found the bivalent BA.1 boosters increased protection to a peak of approximately 53% in addition to the protection present following waning of the monovalent vaccines.

In those aged 65 years and older, higher aVE against hospitalisation was observed than in the younger adults. This may be due to more incidental admissions amongst younger adults which can’t be prevented by vaccination resulting in lower VE estimates in this group (4), and it is likely that the aVE observed amongst those aged 65 years and older is more reflective of the true VE against severe disease. In these analyses the unvaccinated were the comparator group and it is also likely that undocumented prior infections which can’t be adjusted for attenuate VE estimates. Reassuringly, we observed moderate VE of 40-50% at long intervals (over a year) after a second or at least a third dose in the older adults. Since these adults were eligible for the bivalent BA.1 booster, it is possible that some may not have been vaccinated with a bivalent booster vaccine due to a recent infection; this could be biasing these VE estimates up but would likely only affect the early period post-vaccination.

A reduction in VE over time was observed for both the monovalent and bivalent vaccines. This is likely due to waning of effectiveness (25) but could also be due to small changes in effectiveness against the different sub-lineages which emerged during the study period. Previously we observed that VE against BA.2 may have been lower than against BA.1 (3), while VE against severe disease with BA.4/5 was comparable to that observed against BA.2 (2). Early data suggest VE against BQ.1/BQ.1.1 and CH.1.1 may be lower than against BA.5 but numbers of cases were small in this analysis and differences were not statistically significant; this work is ongoing (8).

Data from the Nordic countries and the United States (US) found VE against hospitalisation with bivalent BA.1 and BA.4-5 booster vaccines was high (22, 26-28). A TNCC study from the US found the VE of a BA.4-5 bivalent booster was comparable our estimates; 53% when the last dose was 8-10 months prior (22). Although VE can’t be directly compared between countries, these results suggest there aren’t major differences in the VE of the BA.1 and BA4/5 boosters. We were unable to directly compare the effectiveness of the bivalent and monovalent boosters since these vaccines were not in use during the same period. Other studies have found some evidence that VE of a bivalent booster may be higher than that of the monovalent booster (26, 27). However, when comparing the VE of the bivalent booster to the VE amongst those who received a monovalent fourth dose relative to those with a waned third dose, there was little difference in the iVE observed. As such, there is little evidence that the bivalent BA.1 vaccines conferred increased protection this autumn as compared to the monovalent spring booster although VE may differ slightly due to different circulating sub-lineages. We did not observe any difference by manufacturer in the VE of the bivalent BA.1 boosters.

We estimated the iVE of the bivalent BA.1 boosters compared to those with waned immunity as we considered it more relevant to estimate the additional benefit of the booster since almost all individuals who were vaccinated with the bivalent boosters have had multiple prior doses. Absolute VE can be calculated from the iVE; if the VE amongst those with waned immunity is ~50% and the iVE is ~50% this would translate to an aVE of ~75%. Previous positivity was adjusted for, but many past infections will not be recorded since freely available community testing ended at the end of March 2022. Missing data on past positivity may be expected to bias VE to be lower because past positivity is protective itself and also associated with fewer vaccine doses. Not adjusting for known past positivity made little difference to estimates suggesting this is not likely to lead to a large bias.

The bivalent boosters were protective in the 0 to 6 day interval post vaccination. This is earlier than we would expect to see a true protective effect and could reflect the fact that someone who is unwell and tests positive at home by lateral flow is likely to postpone a vaccination due shortly after, hence leading to fewer positive tests shortly after vaccination (29). This bias should not affect later periods post-vaccination. Overall, our data demonstrate that time since vaccination is more important in determining protection than the total number of doses received. The iVE of the bivalent boosters increased as time since the previous dose increased due to waning of the prior dose-this was also observed in studies from the US (22). Furthermore, there was no difference in the iVE of the bivalent booster if it was a at least a fourth dose relative to the next most recent, or at least a third dose relative to the next most recent. The analyses of the monovalent vaccines also demonstrated the importance of time since vaccination as opposed to dose number in determining VE. After either a second dose or a dose which was at least a third dose, VE was boosted to a similar level before waning to a similar baseline level after approximately one year. For those aged 75 years and older who received a fourth dose, the level of protection waned to the baseline observed after a waned third dose after 6 to 9 months. In spite of waning, these data provide reassuring evidence that a modest level of protection is maintained even in older adults who are more vulnerable to severe disease.

In this study we find reassuring evidence of the long-lasting duration of protection against severe disease-from 6 months after 3 or more doses protection appears to plateau suggesting that further doses may not be needed for those not at high risk of severe disease. Furthermore, we find evidence that bivalent BA.1 boosters vaccines provide additional protection against severe disease in older individuals who are at higher risk of severe disease.

## Data Availability

All data produced in the present work are contained in the manuscript.

## Authors’ contributions

FCMK wrote the manuscript. JLB, NA, FCMK and MR conceptualised the study. FCMK and JS curated the data. FCMK and NA conducted the formal analysis. All co-authors reviewed the manuscript.

## Data Sharing statement

This work is carried out under Regulation 3 of The Health Service (Control of Patient Information) (Secretary of State for Health, 2002) using patient identification information without individual patient consent. Data cannot be made publicly available for ethical and legal reasons, i.e. public availability would compromise patient confidentiality as data tables list single counts of individuals rather than aggregated data.

## Ethics committee approval

UKHSA has legal permission, provided by Regulation 3 of The Health Service (Control of Patient Information) Regulations 2002, to process patient confidential information for national surveillance of communicable diseases and as such, individual patient consent is not required to access records.

