## Supplementary Appendix for "Long-term duration of protection of ancestral-strain monovalent vaccines and effectiveness of the bivalent BA.1 boosters against COVID-19 hospitalisation during a period of BA.5, BQ.1, CH.1.1. and XBB.1.5 circulation in England"

Supplementary Table 1. SUS Acute respiratory illness ICD10 code list.

| SUS Acute respiratory illness ICD10 code list | |
|---|---|
| J04* | Acute laryngitis and tracheitis |
| J09* | Influenza due to identified avian influenza virus |
| J10* | Influenza with pneumonia, other influenza virus identified |
| J11* | Influenza with pneumonia, virus not identified |
| J12* | Viral pneumonia, not elsewhere classified |
| J13* | Pneumonia due to Streptococcus pneumoniae |
| J14* | Pneumonia due to Haemophilus influenzae |
| J15* | Bacterial pneumonia, not elsewhere classified |
| J16* | Pneumonia due to other infectious organisms, not elsewhere classified |
| J17* | Pneumonia in diseases classified elsewhere |
| J18* | Pneumonia, organism unspecified |
| J20* | Acute bronchitis |
| J21* | Acute bronchiolitis |
| J22* | Unspecified acute lower respiratory infection |
| J80* | ARDS (related to respiratory infection) |
| U071, U072 | COVID-19, virus identified and not identified |
| U04* | Severe acute respiratory syndrome (SARS) |

Supplementary Table 2. Descriptive characteristics of those included the absolute vaccine effectiveness (aVE) analysis of the duration of protection of the monovalent vaccines.

| | | | Overall | | Controls | | Cases | |
|---|---|---|---|---|---|---|---|---|
| | | | n | % | n | % | n | % |
| Vaccination Status | No. of doses | Interval since last dose | 63,251 | 100% | 43,410 | 69% | 19,841 | 31% |
| | Unvaccinated | n/a | 4,848 | 7.7% | 2,805 | 6.5% | 2,043 | 10.3% |
| | 2 | 0-13 days | 1 | 0.0% | 0 | 0.0% | 1 | 0.0% |
| | | 2 -11 weeks | 59 | 0.1% | 44 | 0.1% | 15 | 0.1% |
| | | 3-5 months | 191 | 0.3% | 150 | 0.3% | 41 | 0.2% |
| | | 6 - 8 months | 387 | 0.6% | 266 | 0.6% | 121 | 0.6% |
| | | 9- 11 months | 652 | 1.0% | 448 | 1.0% | 204 | 1.0% |
| | | 12 - 14 months | 1,682 | 2.7% | 1,039 | 2.4% | 643 | 3.2% |
| | | 15+ months | 2,674 | 4.2% | 1,913 | 4.4% | 761 | 3.8% |
| | 3+ | 0-13 days | 647 | 1.0% | 474 | 1.1% | 173 | 0.9% |
| | | 2 -11 weeks | 10,500 | 16.6% | 7,848 | 18.1% | 2,652 | 13.4% |
| | | 3-5 months | 14,874 | 23.5% | 10,409 | 24.0% | 4,465 | 22.5% |
| | | 6 - 8 months | 14,459 | 22.9% | 9,143 | 21.1% | 5,316 | 26.8% |
| | | 9- 11 months | 10,240 | 16.2% | 7,247 | 16.7% | 2,993 | 15.1% |
| | | 12 - 14 months | 2,035 | 3.2% | 1,624 | 3.7% | 411 | 2.1% |
| | | 15+ months | 2 | 0.0% | 0 | 0.0% | 2 | 0.0% |
| Gender | Female | | 31,955 | 50.5% | 22,083 | 50.9% | 9,872 | 49.8% |
| | Male | | 30,846 | 48.8% | 20,923 | 48.2% | 9,923 | 50.0% |
| | Missing | | 450 | 0.7% | 404 | 0.9% | 46 | 0.2% |
| Age | 18-19 | | 207 | 0.3% | 154 | 0.4% | 53 | 0.3% |
| | 20-24 | | 572 | 0.9% | 398 | 0.9% | 174 | 0.9% |
| | 25-29 | | 747 | 1.2% | 481 | 1.1% | 266 | 1.3% |
| | 30-34 | | 949 | 1.5% | 668 | 1.5% | 281 | 1.4% |
| | 35-39 | | 1,072 | 1.7% | 804 | 1.9% | 268 | 1.4% |
| | 40-44 | | 1,256 | 2.0% | 970 | 2.2% | 286 | 1.4% |
| | 45-49 | | 1,518 | 2.4% | 1,116 | 2.6% | 402 | 2.0% |
| | 50-54 | | 2,339 | 3.7% | 1,717 | 4.0% | 622 | 3.1% |
| | 55-59 | | 3,192 | 5.0% | 2,331 | 5.4% | 861 | 4.3% |
| | 60-64 | | 4,210 | 6.7% | 3,050 | 7.0% | 1,160 | 5.8% |
| | 65-69 | | 5,149 | 8.1% | 3,505 | 8.1% | 1,644 | 8.3% |
| | 70-74 | | 7,183 | 11.4% | 4,721 | 10.9% | 2,462 | 12.4% |
| | 75-79 | | 8,875 | 14.0% | 6,109 | 14.1% | 2,766 | 13.9% |
| | 80-84 | | 9,253 | 14.6% | 6,207 | 14.3% | 3,046 | 15.4% |
| | 85-89 | | 9,077 | 14.4% | 5,974 | 13.8% | 3,103 | 15.6% |
| | 90+ | | 7,652 | 12.1% | 5,205 | 12.0% | 2,447 | 12.3% |
| Ethnicity | African | | 561 | 0.9% | 380 | 0.9% | 181 | 0.9% |
| | Any other Asian background | | 637 | 1.0% | 435 | 1.0% | 202 | 1.0% |
| | Any other Black background | | 279 | 0.4% | 190 | 0.4% | 89 | 0.4% |
| | Any other White background | | 3,194 | 5.0% | 2,143 | 4.9% | 1,051 | 5.3% |
| | Any other ethnic group | | 758 | 1.2% | 518 | 1.2% | 240 | 1.2% |

| | | | | | | | |
|---|---|---|---|---|---|---|---|
| | Any other mixed background | 251 | 0.4% | 171 | 0.4% | 80 | 0.4% |
| | Bangladeshi or British Bangladeshi | 327 | 0.5% | 225 | 0.5% | 102 | 0.5% |
| | British, Mixed British | 50,773 | 80.3% | 34,969 | 80.6% | 15,804 | 79.7% |
| | Caribbean | 634 | 1.0% | 392 | 0.9% | 242 | 1.2% |
| | Chinese | 119 | 0.2% | 74 | 0.2% | 45 | 0.2% |
| | Indian or British Indian | 1,335 | 2.1% | 886 | 2.0% | 449 | 2.3% |
| | Irish | 754 | 1.2% | 521 | 1.2% | 233 | 1.2% |
| | Pakistani or British Pakistani | 1,082 | 1.7% | 763 | 1.8% | 319 | 1.6% |
| | White and Asian | 67 | 0.1% | 43 | 0.1% | 24 | 0.1% |
| | White and Black African | 54 | 0.1% | 32 | 0.1% | 22 | 0.1% |
| | White and Black Caribbean | 124 | 0.2% | 84 | 0.2% | 40 | 0.2% |
| | Missing | 2,302 | 3.6% | 1,584 | 3.6% | 718 | 3.6% |
| NHS Region | East of England | 5,716 | 9.0% | 4,033 | 9.3% | 1,683 | 8.5% |
| | London | 8,243 | 13.0% | 5,466 | 12.6% | 2,777 | 14.0% |
| | Midlands | 13,698 | 21.7% | 9,353 | 21.5% | 4,345 | 21.9% |
| | North East | 11,407 | 18.0% | 7,611 | 17.5% | 3,796 | 19.1% |
| | North West | 9,958 | 15.7% | 7,168 | 16.5% | 2,790 | 14.1% |
| | South East | 8,027 | 12.7% | 5,614 | 12.9% | 2,413 | 12.2% |
| | South West | 6,202 | 9.8% | 4,165 | 9.6% | 2,037 | 10.3% |
| IMD Quintiles | 1 | 15,723 | 24.9% | 10,872 | 25.0% | 4,851 | 24.4% |
| | 2 | 13,187 | 20.8% | 9,048 | 20.8% | 4,139 | 20.9% |
| | 3 | 12,254 | 19.4% | 8,385 | 19.3% | 3,869 | 19.5% |
| | 4 | 11,731 | 18.5% | 8,030 | 18.5% | 3,701 | 18.7% |
| | 5 | 10,057 | 15.9% | 6,873 | 15.8% | 3,184 | 16.0% |
| | Missing | 299 | 0.5% | 202 | 0.5% | 97 | 0.5% |
| Previously positive | None | 50,553 | 79.9% | 32,558 | 75.0% | 17,995 | 90.7% |
| | Wild-type | 1,622 | 2.6% | 1,324 | 3.0% | 298 | 1.5% |
| | Alpha | 1,981 | 3.1% | 1,590 | 3.7% | 391 | 2.0% |
| | Delta | 2,094 | 3.3% | 1,710 | 3.9% | 384 | 1.9% |
| | Omicron (before 1st April 2022) | 5,347 | 8.5% | 4,711 | 10.9% | 636 | 3.2% |
| | Omicron (from 1st April 2022 onwards) | 1,654 | 2.6% | 1,517 | 3.5% | 137 | 0.7% |
| Risk status | Healthcare worker | 555 | 0.9% | 383 | 0.9% | 172 | 0.9% |
| | Carehome | 2,460 | 3.9% | 1,957 | 4.5% | 503 | 2.5% |
| | CaaS autumn booster cohort | 46,825 | 74.0% | 31,271 | 72.0% | 15,554 | 78.4% |
| | At risk - ever, from NIMS* | 12,165 | 19.2% | 8,705 | 20.1% | 3,460 | 17.4% |
| | CEV | 29,167 | 46.1% | 20,005 | 46.1% | 9,162 | 46.2% |
| | Severely immunosuppressed | 8,074 | 12.8% | 5,517 | 12.7% | 2,557 | 12.9% |

*Only for those <65 years

Supplementary Figure 1. Cases (those testing positive) and controls (those testing negative) over time in the analysis of duration of protection of the monovalent vaccines.

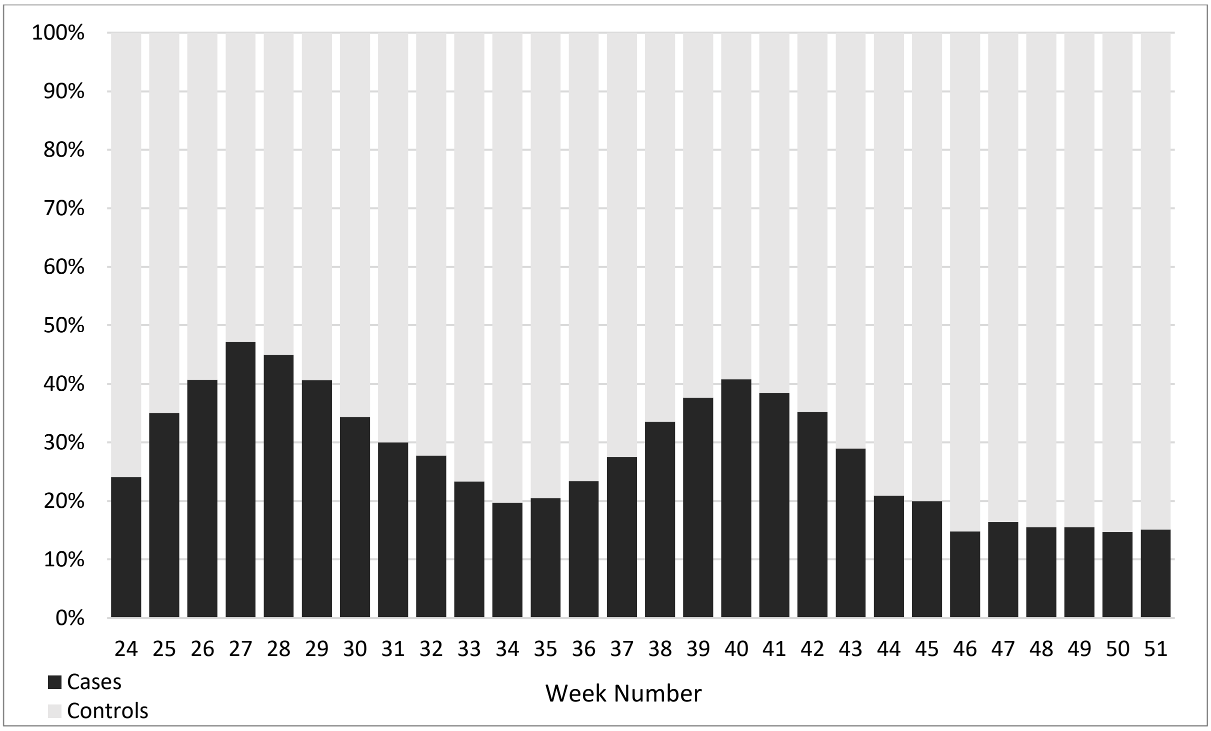

Supplementary Table 3. Absolute VE (aVE) against hospitalisation amongst those aged 65 years and older, without the adjustment for previous positivity.

| Age | No. of doses | Interval since last dose | Controls | Cases | OR | VE |
|---|---|---|---|---|---|---|
| 65+ | Unvaccinated | | 1172 | 685 | Baseline | Baseline |
| | 2 | 0-13 days | 0 | 0 | n too small | n too small |
| | | 2 -11 weeks | 4 | 0 | n too small | n too small |
| | | 3-5 months | 26 | 6 | n too small | n too small |
| | | 6 - 8 months | 33 | 10 | 0.45 (0.21-0.93) | 55.4 (6.9 to 78.6) |
| | | 9- 11 months | 80 | 28 | 0.51 (0.32-0.81) | 48.8 (19.0 to 67.6) |
| | | 12 - 14 months | 99 | 33 | 0.61 (0.4-0.93) | 39.3 (7.3 to 60.3) |
| | | 15+ months | 1282 | 504 | 0.66 (0.57-0.77) | 33.9 (23.5 to 42.8) |
| | 3+ | 0-13 days | 3 | 0 | n too small | n too small |
| | | 2 -11 weeks | 647 | 183 | 0.29 (0.24-0.36) | 70.6 (64.0 to 76.0) |
| | | 3-5 months | 4598 | 2051 | 0.45 (0.4-0.5) | 55.3 (49.5 to 60.3) |
| | | 6 - 8 months | 2639 | 1168 | 0.6 (0.53-0.68) | 40.3 (32.4 to 47.3) |
| | | 9- 11 months | 3178 | 1675 | 0.64 (0.56-0.72) | 36.4 (28.3 to 43.7) |
| | | 12 - 14 months | 2603 | 799 | 0.62 (0.55-0.71) | 37.6 (29.0 to 45.2) |
| | | 15+ months | 91 | 34 | 0.72 (0.48-1.09) | 27.8 (-9.4 to 52.4) |

Supplementary Table 4. Incremental vaccine effectiveness (iVE) against hospitalisation of at least 4 doses (monovalent vaccines) in individuals aged 75 years and older.

| No. of doses | Interval since last dose | Controls | Cases | OR | iVE |
|---|---|---|---|---|---|
| 3 | 6+ months | 5039 | 3004 | Baseline | Baseline |
| 4+ | 0 - 13 days | 329 | 125 | 0.53 (0.43-0.67) | 46.7 (33.4 to 57.4) |
| | 2 - 11 weeks | 6499 | 2221 | 0.48 (0.45-0.52) | 51.9 (48.2 to 55.3) |
| | 3 - 5 months | 8103 | 3572 | 0.73 (0.68-0.78) | 26.8 (21.7 to 31.6) |
| | 6 - 9 months | 1208 | 653 | 1.02 (0.9-1.15) | -1.8 (-15.1 to 10.0) |

Supplementary Table 5. Descriptive characteristics of those included the incremental vaccine effectiveness (iVE) analysis of the bivalent BA.1 booster vaccine.

| | | | Overall | | Controls | | Cases | |
|---|---|---|---|---|---|---|---|---|
| | | | n | % | n | % | n | % |
| Vaccination Status | Bivalent vaccine | Interval | 49,062 | 100% | 39,108 | 80% | 9,954 | 20% |
| | None | | 19,108 | 38.9% | 13,945 | 35.7% | 5,163 | 51.9% |
| | Pfizer | 0-6 days | 408 | 0.8% | 303 | 0.8% | 105 | 1.1% |
| | | 7-13 days | 581 | 1.2% | 463 | 1.2% | 118 | 1.2% |
| | | 2-4 weeks | 1,917 | 3.9% | 1,649 | 4.2% | 268 | 2.7% |
| | | 5-9 weeks | 5,072 | 10.3% | 4,394 | 11.2% | 678 | 6.8% |
| | | 10+ weeks | 3,661 | 7.5% | 3,086 | 7.9% | 575 | 5.8% |
| | Moderna | 0-6 days | 279 | 0.6% | 195 | 0.5% | 84 | 0.8% |
| | | 7-13 days | 369 | 0.8% | 269 | 0.7% | 100 | 1.0% |
| | | 2-4 weeks | 1,462 | 3.0% | 1,156 | 3.0% | 306 | 3.1% |
| | | 5-9 weeks | 4,404 | 9.0% | 3,760 | 9.6% | 644 | 6.5% |
| | | 10+ weeks | 11,801 | 24.1% | 9,888 | 25.3% | 1,913 | 19.2% |
| Gender | Female | | 25,520 | 52.0% | 20,656 | 52.8% | 4,864 | 48.9% |
| | Male | | 22,990 | 46.9% | 17,927 | 45.8% | 5,063 | 50.9% |
| | Missing | | 552 | 1.1% | 525 | 1.3% | 27 | 0.3% |
| Age | 50-54 | | 1,968 | 4.0% | 1,655 | 4.2% | 313 | 3.1% |
| | 55-59 | | 2,854 | 5.8% | 2,405 | 6.1% | 449 | 4.5% |
| | 60-64 | | 3,967 | 8.1% | 3,326 | 8.5% | 641 | 6.4% |
| | 65-69 | | 4,954 | 10.1% | 4,042 | 10.3% | 912 | 9.2% |
| | 70-74 | | 6,768 | 13.8% | 5,381 | 13.8% | 1,387 | 13.9% |
| | 75-79 | | 7,524 | 15.3% | 6,006 | 15.4% | 1,518 | 15.3% |
| | 80-84 | | 7,793 | 15.9% | 6,009 | 15.4% | 1,784 | 17.9% |
| | 85-89 | | 7,358 | 15.0% | 5,694 | 14.6% | 1,664 | 16.7% |
| | 90+ | | 5,876 | 12.0% | 4,590 | 11.7% | 1,286 | 12.9% |
| Ethnicity | African | | 275 | 0.6% | 213 | 0.5% | 62 | 0.6% |
| | Any other Asian background | | 410 | 0.8% | 330 | 0.8% | 80 | 0.8% |
| | Any other Black background | | 130 | 0.3% | 101 | 0.3% | 29 | 0.3% |
| | Any other White background | | 2,180 | 4.4% | 1,733 | 4.4% | 447 | 4.5% |
| | Any other ethnic group | | 455 | 0.9% | 370 | 0.9% | 85 | 0.9% |
| | Any other mixed background | | 154 | 0.3% | 127 | 0.3% | 27 | 0.3% |
| | Bangladeshi or British Bangladeshi | | 205 | 0.4% | 175 | 0.4% | 30 | 0.3% |
| | British, Mixed British | | 40,761 | 83.1% | 32,420 | 82.9% | 8,341 | 83.8% |
| | Caribbean | | 347 | 0.7% | 272 | 0.7% | 75 | 0.8% |
| | Chinese | | 65 | 0.1% | 53 | 0.1% | 12 | 0.1% |
| | Indian or British Indian | | 996 | 2.0% | 816 | 2.1% | 180 | 1.8% |
| | Irish | | 581 | 1.2% | 477 | 1.2% | 104 | 1.0% |
| | Pakistani or British Pakistani | | 630 | 1.3% | 538 | 1.4% | 92 | 0.9% |
| | White and Asian | | 42 | 0.1% | 32 | 0.1% | 10 | 0.1% |
| | White and Black African | | 41 | 0.1% | 32 | 0.1% | 9 | 0.1% |
| | White and Black Caribbean | | 65 | 0.1% | 49 | 0.1% | 16 | 0.2% |

| | | | | | | | |
|---|---|---|---|---|---|---|---|
| | Missing | 1,725 | 3.5% | 1,370 | 3.5% | 355 | 3.6% |
| NHS Region | East of England | 3,830 | 7.8% | 2,985 | 7.6% | 845 | 8.5% |
| | London | 5,874 | 12.0% | 4,728 | 12.1% | 1,146 | 11.5% |
| | Midlands | 10,334 | 21.1% | 8,126 | 20.8% | 2,208 | 22.2% |
| | North East | 10,092 | 20.6% | 8,051 | 20.6% | 2,041 | 20.5% |
| | North West | 7,488 | 15.3% | 6,263 | 16.0% | 1,225 | 12.3% |
| | South East | 5,949 | 12.1% | 4,633 | 11.8% | 1,316 | 13.2% |
| | South West | 5,495 | 11.2% | 4,322 | 11.1% | 1,173 | 11.8% |
| IMD Quintiles | 1 | 11,493 | 23.4% | 9,247 | 23.6% | 2,246 | 22.6% |
| | 2 | 9,925 | 20.2% | 7,931 | 20.3% | 1,994 | 20.0% |
| | 3 | 9,786 | 19.9% | 7,834 | 20.0% | 1,952 | 19.6% |
| | 4 | 9,325 | 19.0% | 7,382 | 18.9% | 1,943 | 19.5% |
| | 5 | 8,386 | 17.1% | 6,594 | 16.9% | 1,792 | 18.0% |
| | Missing | 147 | 0.3% | 120 | 0.3% | 27 | 0.3% |
| Previously positive | None | 37,497 | 76.4% | 28,862 | 73.8% | 8,635 | 86.7% |
| | Wild-type | 1,203 | 2.5% | 1,042 | 2.7% | 161 | 1.6% |
| | Alpha | 1,444 | 2.9% | 1,238 | 3.2% | 206 | 2.1% |
| | Delta | 1,504 | 3.1% | 1,305 | 3.3% | 199 | 2.0% |
| | Omicron (before 1st April 2022) | 4,276 | 8.7% | 3,778 | 9.7% | 498 | 5.0% |
| | Omicron (from 1st April 2022 onwards) | 3,138 | 6.4% | 2,883 | 7.4% | 255 | 2.6% |
| Risk status | Healthcare worker | 316 | 0.6% | 259 | 0.7% | 57 | 0.6% |
| | Carehome | 2,178 | 4.4% | 1,839 | 4.7% | 339 | 3.4% |
| | CaaS autumn booster cohort | 44,002 | 89.7% | 34,842 | 89.1% | 9,160 | 92.0% |
| | At risk - ever, from NIMS* | 7,615 | 15.5% | 6,284 | 16.1% | 1,331 | 13.4% |
| | CEV | 21,920 | 44.7% | 17,276 | 44.2% | 4,644 | 46.7% |
| | Severely immunosuppressed | 4,986 | 10.2% | 3,727 | 9.5% | 1,259 | 12.6% |

*Only for those <65 years

Supplementary Figure 2. Cases (those testing positive) and controls (those testing negative) over time in the bivalent BA.1 booster incremental VE (iVE) analysis.

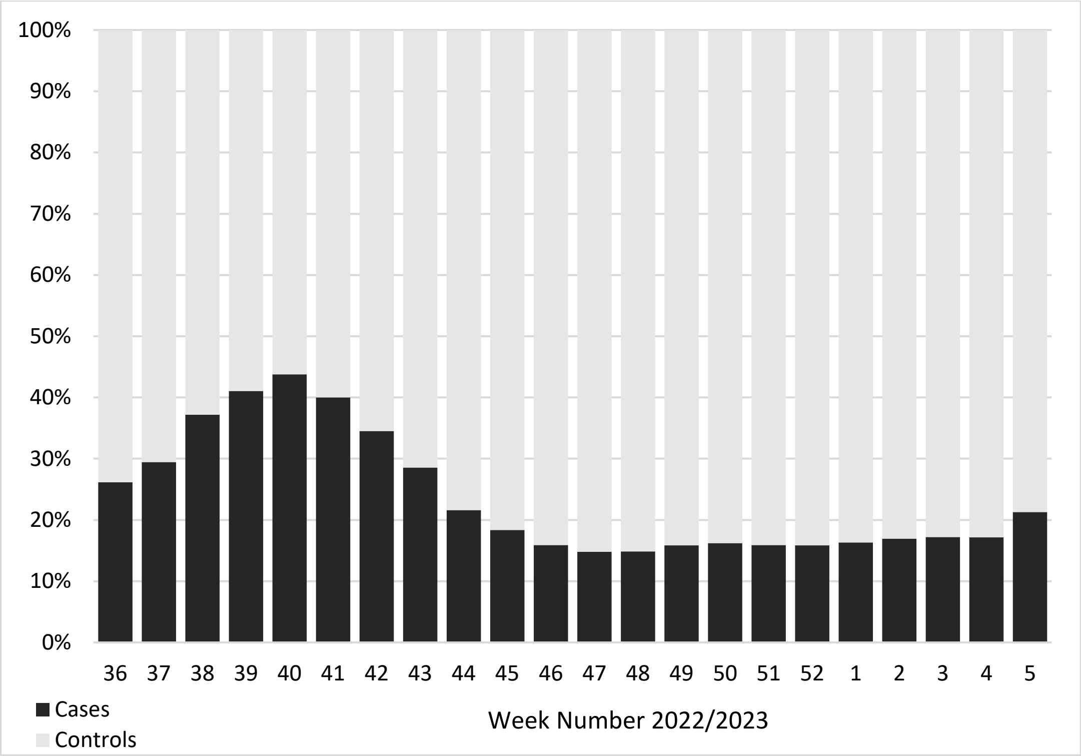

Supplementary Figure 3. Distribution of time in days since vaccination amongst those who received a bivalent booster.

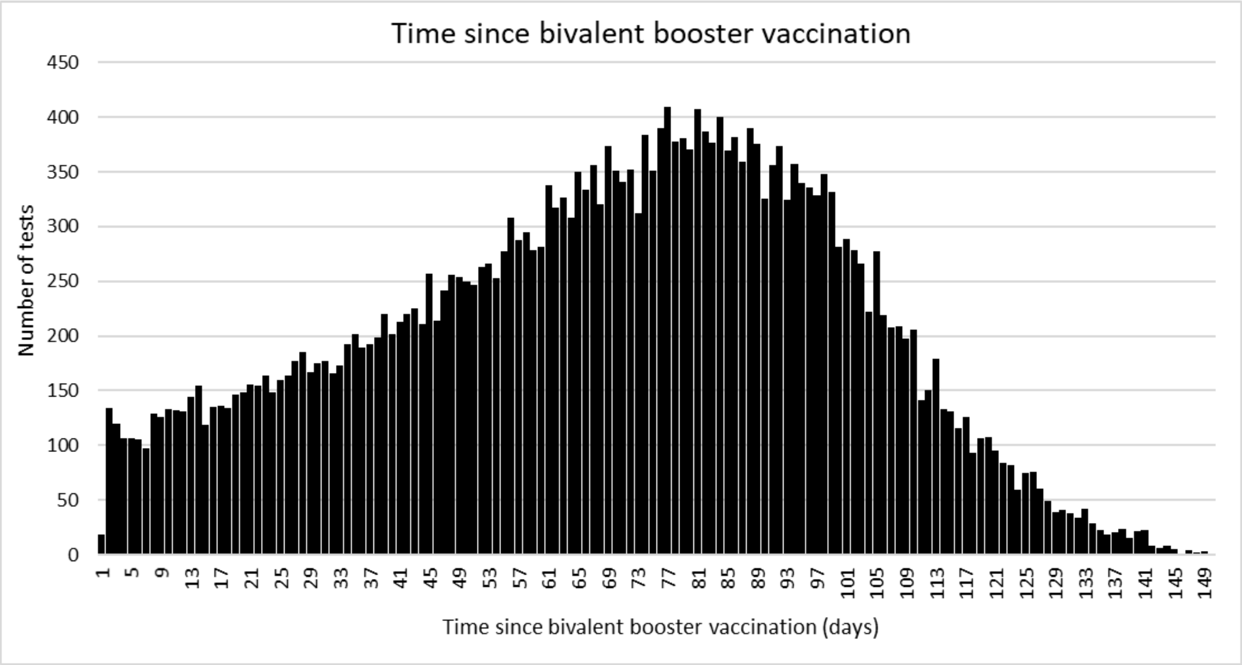

Supplementary Table 6. Stratified and sensitivity analyses of incremental vaccine effectiveness (iVE) of the bivalent BA.1 booster vaccines against hospitalisation.

| Bivalent vaccine | Interval (weeks) | Controls | Cases | Odds Ratio | iVE (95% C.I.) |
|---|---|---|---|---|---|
| 2+ doses, with a 6 week to 5 month interval to the most recent dose | | | | | |
| None | - | 4430 | 2058 | Baseline | Baseline |
| Pfizer or Moderna | 0-6 days | 425 | 142 | 0.69 (0.56-0.85) | 30.5 (14.6 to 43.5) |
| | 7-13 days | 431 | 176 | 0.76 (0.62-0.92) | 24.2 (8.0 to 37.5) |
| | 2-4 weeks | 1128 | 269 | 0.54 (0.46-0.63) | 46.0 (36.8 to 53.8) |
| | 5-9 weeks | 1044 | 163 | 0.56 (0.46-0.68) | 44.2 (31.7 to 54.4) |
| | 10+ weeks | 174 | 26 | 0.54 (0.34-0.85) | 46.3 (14.7 to 66.2) |
| 2+ doses, with a 6 to 8 month interval to the most recent dose | | | | | |
| None | - | 2903 | 1251 | Baseline | Baseline |
| Pfizer or Moderna | 0-6 days | 195 | 72 | 0.6 (0.44-0.8) | 40.3 (19.7 to 55.6) |
| | 7-13 days | 266 | 115 | 0.75 (0.59-0.95) | 25.3 (4.6 to 41.5) |
| | 2-4 weeks | 1329 | 348 | 0.54 (0.46-0.62) | 46.5 (37.9 to 53.9) |
| | 5-9 weeks | 4489 | 880 | 0.61 (0.54-0.68) | 39.2 (31.8 to 45.8) |
| | 10+ weeks | 7221 | 1484 | 0.66 (0.59-0.74) | 33.7 (25.7 to 40.8) |
| 2+ doses, with a 9 to 11 month interval to the most recent dose | | | | | |
| None | - | 4863 | 2152 | Baseline | Baseline |
| Pfizer or Moderna | 0-6 days | 222 | 90 | 0.7 (0.54-0.91) | 29.7 (8.6 to 45.9) |
| | 7-13 days | 302 | 69 | 0.44 (0.33-0.58) | 55.9 (41.8 to 66.7) |
| | 2-4 weeks | 710 | 124 | 0.4 (0.33-0.5) | 59.9 (50.5 to 67.5) |
| | 5-9 weeks | 830 | 145 | 0.62 (0.51-0.77) | 37.5 (22.9 to 49.3) |
| | 10+ weeks | 2067 | 458 | 0.76 (0.61-0.94) | 24.5 (5.9 to 39.4) |
| 2+ doses, with at least a 12 month interval to the most recent dose | | | | | |
| None | - | 6179 | 1760 | Baseline | Baseline |
| Pfizer or Moderna | 0-6 days | 81 | 27 | 0.8 (0.5-1.28) | 20.0 (-28 to 49.9) |
| | 7-13 days | 164 | 34 | 0.61 (0.42-0.9) | 38.7 (9.5 to 58.4) |
| | 2-4 weeks | 766 | 102 | 0.46 (0.37-0.58) | 53.7 (42.1 to 62.9) |
| | 5-9 weeks | 2835 | 297 | 0.45 (0.39-0.51) | 55.4 (48.7 to 61.2) |
| | 10+ weeks | 3686 | 546 | 0.62 (0.55-0.7) | 38.1 (30.2 to 45.0) |
| 2+ doses, with at least a 6 month interval to the most recent dose, all hospital admissions | | | | | |
| None | - | 92863 | 11271 | Baseline | Baseline |
| Pfizer or Moderna | 0-6 days | 3480 | 355 | 0.64 (0.57-0.71) | 36.4 (28.7 to 43.2) |
| | 7-13 days | 4766 | 459 | 0.61 (0.55-0.67) | 39.5 (33.1 to 45.2) |
| | 2-4 weeks | 18206 | 1222 | 0.44 (0.42-0.47) | 55.5 (52.5 to 58.3) |
| | 5-9 weeks | 39474 | 2939 | 0.53 (0.51-0.56) | 46.7 (44.1 to 49.2) |
| | 10+ weeks | 60294 | 6158 | 0.66 (0.63-0.69) | 34.3 (31.4 to 37.1) |
| 2+ doses, with at least a 6 month interval to the most recent dose, no adjustment for previous positivity | | | | | |
| None | - | 13945 | 5163 | Baseline | Baseline |
| Pfizer or Moderna | 0-6 days | 498 | 189 | 0.66 (0.55-0.78) | 34.4 (21.6 to 45.1) |
| | 7-13 days | 732 | 218 | 0.58 (0.49-0.68) | 42.4 (32.3 to 51.0) |
| | 2-4 weeks | 2805 | 574 | 0.47 (0.43-0.52) | 52.9 (47.9 to 57.5) |
| | 5-9 weeks | 8154 | 1322 | 0.54 (0.5-0.58) | 45.7 (41.5 to 49.7) |

| | | | | | |
|---|---|---|---|---|---|
| | 10+ weeks | 12974 | 2488 | 0.64 (0.6-0.69) | 35.9 (31.3 to 40.2) |
| 3+ doses, with at least a 6 month interval to the most recent dose | | | | | |
| None | - | 11499 | 4396 | Baseline | Baseline |
| Pfizer or Moderna | 0-6 days | 473 | 184 | 0.68 (0.57-0.82) | 31.8 (18.1 to 43.2) |
| | 7-13 days | 711 | 213 | 0.59 (0.5-0.7) | 40.7 (30.0 to 49.7) |
| | 2-4 weeks | 2735 | 561 | 0.48 (0.43-0.53) | 52.4 (47.1 to 57.1) |
| | 5-9 weeks | 8026 | 1298 | 0.55 (0.5-0.59) | 45.4 (41.0 to 49.5) |
| | 10+ weeks | 12827 | 2473 | 0.65 (0.6-0.7) | 35.2 (30.2 to 39.8) |
| 4+ doses, with at least a 6 month interval to the most recent dose | | | | | |
| None | - | 2511 | 1110 | Baseline | Baseline |
| Pfizer or Moderna | 0-6 days | 181 | 74 | 0.63 (0.47-0.85) | 37.3 (15.4 to 53.5) |
| | 7-13 days | 264 | 115 | 0.75 (0.59-0.96) | 24.9 (3.9 to 41.3) |
| | 2-4 weeks | 1336 | 353 | 0.53 (0.45-0.61) | 47.3 (38.9 to 54.6) |
| | 5-9 weeks | 4704 | 913 | 0.59 (0.53-0.66) | 40.9 (33.8 to 47.2) |
| | 10+ weeks | 8993 | 1887 | 0.67 (0.6-0.74) | 33.5 (25.7 to 40.4) |
| 4+ doses, with at a 6 to 8 month interval to the most recent dose | | | | | |
| None | - | 2280 | 1038 | Baseline | Baseline |
| Pfizer or Moderna | 0-6 days | 175 | 69 | 0.6 (0.44-0.81) | 40.1 (18.6 to 56.0) |
| | 7-13 days | 252 | 110 | 0.73 (0.57-0.94) | 27.0 (6.1 to 43.3) |
| | 2-4 weeks | 1283 | 339 | 0.52 (0.45-0.6) | 48.1 (39.5 to 55.4) |
| | 5-9 weeks | 4381 | 850 | 0.59 (0.52-0.66) | 41.1 (33.7 to 47.7) |
| | 10+ weeks | 7075 | 1459 | 0.66 (0.58-0.74) | 34.3 (26.1 to 41.6) |
| Age 75 years+, 4+ doses, with at a 6 to 8 month interval to the most recent dose | | | | | |
| None | - | 1931 | 882 | Baseline | Baseline |
| Pfizer or Moderna | 0-6 days | 151 | 61 | 0.59 (0.42-0.82) | 41.0 (17.9 to 57.6) |
| | 7-13 days | 229 | 97 | 0.69 (0.53-0.9) | 31.3 (10.2 to 47.5) |
| | 2-4 weeks | 1135 | 311 | 0.52 (0.44-0.61) | 48.0 (38.8 to 55.7) |
| | 5-9 weeks | 3937 | 784 | 0.6 (0.53-0.68) | 40.0 (32.0 to 47.0) |
| | 10+ weeks | 6333 | 1313 | 0.66 (0.58-0.75) | 33.8 (25.0 to 41.5) |
| Age 65+, 2+ doses, with at least a 6 month interval to the most recent dose | | | | | |
| None | - | 9531 | 4111 | Baseline | Baseline |
| Pfizer or Moderna | 0-6 days | 403 | 164 | 0.64 (0.53-0.78) | 36.0 (22.2 to 47.3) |
| | 7-13 days | 575 | 192 | 0.58 (0.49-0.69) | 42.0 (30.8 to 51.4) |
| | 2-4 weeks | 2289 | 507 | 0.45 (0.4-0.5) | 55.2 (50.0 to 59.9) |
| | 5-9 weeks | 6865 | 1203 | 0.53 (0.49-0.58) | 47.0 (42.5 to 51.1) |
| | 10+ weeks | 11741 | 2311 | 0.62 (0.58-0.67) | 37.7 (32.8 to 42.2) |
